# Predicting the efficacy of Ervebo against Bundibugyo virus disease

**DOI:** 10.64898/2026.10.02.26364627

**Authors:** Karen M Elias, Ece Egilmezer, Shanchita R Khan, Ainslie Mitchell, Arnold Reynaldi, Matthew T Berry, Timothy E Schlub, Bronwyn A Bailey, Deborah Cromer, Tari Turner, Eva Stadler, Miles P Davenport, David S Khoury

## Abstract

There is currently no clinical data for the effectiveness of the only licensed Ebola virus disease vaccine (Ervebo) against Bundibugyo virus disease. A number of studies have shown immune cross-reactivity with Bundibugyo virus after Ervebo vaccination. However, antibody recognition of Bundibugyo is around 2.8-fold lower than recognition of Ebola virus. Here, we aimed to infer how a 2.8-fold drop in immune recognition may affect Ervebo protection against Bundibugyo virus disease based on analysis of data on Ervebo immunogenicity and protection from Ebola virus disease. This work has three main components. Firstly, we analysed the timing of vaccine protection in the Ervebo pivotal ring vaccination clinical trial. Secondly, we performed a systematic review and meta-analysis of antibody responses to Ebola virus after Ervebo vaccination over time. Thirdly, we combined these data with previously reported data on the cross-reactivity to Bundibugyo virus to infer protection of Ervebo against Bundibugyo virus disease.

We find that the clinical trial data for Ervebo supports vaccine protection beginning earlier than 10 days after vaccination. Secondly, binding antibody levels against Ebola virus on day 28 post vaccination (peak responses) were on average 12.9-fold (95% confidence interval, CI: 6.4 – 26.0) higher than on day 7, 6.3-fold (95% CI: 3.7 – 10.8) higher than on day 10, and 2.5-fold (95% CI: 1.4 – 4.4) higher than on day 14 (neutralising antibodies showed similar results). Finally, we consider the scenarios where antibody levels to Ebola virus at day 7 or 10 are putative protective thresholds against Ebola virus disease, and we assume these thresholds also apply for Bundibugyo virus disease. Then, with a 2.8-fold reduction of responses to Bundibugyo compared to Ebola virus, we predict antibody responses to Bundibugyo virus after Ervebo vaccination are likely above these putative protective thresholds.

## Introduction

The Bundibugyo virus disease (BVD) outbreak in The Democratic Republic of the Congo was declared a Public Health Emergency of International Concern by the World Health Organization on 17 May 2026^1^. There are currently no licensed therapeutics or vaccines for treating or preventing Bundibugyo virus disease. However, there is an existing licensed vaccine for prevention of Ebola virus disease (EVD) that incorporates Ebola virus (EBOV) glycoprotein (Ervebo, rVSV-EBOV-GP). There is around 65% amino acid similarity between the EBOV and Bundibugyo virus (BDBV) glycoproteins (GP)^2^. A study of monoclonal antibodies from an Ebola survivor suggested that the majority of monoclonals that bound to EBOV glycoprotein (EBOV-GP) also bound to BDBV-GP and around half of the EBOV-GP-specific neutralising antibodies could also neutralise BDBV in vitro (5/9 monoclonals)^3^. A recent meta-analysis suggests that after Ervebo vaccination, both binding and neutralising antibody recognition of BDBV is around 2.8-fold lower than recognition of EBOV^4^. Evidence for antibody cross-reactivity with BDBV after Ervebo vaccination has been discussed both by the WHO and Africa CDC in considering whether to deploy existing stockpiles of Ervebo^5,6^. Ervebo has subsequently been released from the stockpile for use in a ring vaccination randomized controlled trial (RCT) and for expanded vaccination of health care workers. While there is evidence of antibody cross-reactivity to BDBV after Ervebo vaccination, it is unclear whether the level of cross-reactivity is sufficient to provide clinical cross-protection against Bundibugyo virus disease.

The efficacy of Ervebo was determined in a cluster randomized trial assessing the benefits of immediate versus delayed Ervebo vaccination in preventing symptomatic Ebola infection in contacts and contacts of contacts of an index Ebola case. An interim analysis of infection after vaccination showed that no new cases occurred in vaccinated individuals beyond day 6 post vaccination^7^. However, a later, larger analysis from the same study showed no new cases beyond day 9 in vaccinated subjects compared with ongoing infection in the delayed vaccination clusters^8^.

A previous post-hoc analysis of antibody levels 14 days after Ervebo vaccination attempted to infer a ‘protective threshold’ of antibodies required for protection from Ebola infection^9^, based on the observation that protection was achieved by this time. A key assumption of such analyses is that protection from Ebola was achieved early after Ervebo vaccination and that antibody levels at that time are both associated with this protection and predictive of protection at later timepoints.

To predict Ervebo protection against Bundibugyo virus infection, we first re-analysed data from the cluster randomised trial of efficacy of Ervebo to estimate the time at which protection against Ebola virus disease began^8^. We then performed a systematic review to identify studies reporting antibody levels after Ervebo vaccination and used these data to model EBOV-specific antibody levels over time. The estimated EBOV-specific antibody level at the onset of protection was used as a putative protective threshold against Ebola virus disease. Finally, we accounted for the estimated 2.8-fold reduction^4^ in antibody recognition of BDBV relative to EBOV and compared the predicted BDBV-specific antibody levels over time with the putative protection threshold. Our approach provides an indirect means of inferring the efficacy of Ervebo against Bundibugyo virus disease but is highly dependent on several critical assumptions.

## Methods

This study has three main components: (i) a post-hoc analysis of clinical vaccine efficacy data from the pivotal RCT of Ervebo, (ii) a systematic review and meta-analysis of EBOV-specific antibody levels over time in Ervebo-vaccinated individuals (comparing antibody levels between day 7 and 14 with levels after day 21), and (iii) an inference of whether BDBV-specific antibodies are likely to be above a putative protective antibody level based on the previously observed levels of antibody cross-reactivity.

### Extraction of data on Ervebo efficacy against Ebola infection

Only one RCT has been performed to determine the vaccine efficacy of Ervebo against Ebola virus disease, with outcomes published both in an interim and a final report^7,8^. Data on vaccine efficacy from this trial could be derived from the survival curve shown in Figure 3 (top right panel) of the final report^8^. Two authors (DSK and AR) independently extracted the timing of EVD events from this survival curve using WebPlotDigitizer (version v5)^10^. There were no conflicts in the extracted timing of event data. As a quality control on the extracted data, we also cross-checked the numbers of events against those reported in Table 4 of the final report^8^. Ring identifiers were unavailable for extraction, so within-ring clustering could not be accounted for in the analysis. However, the original analysis reported a relatively low intraclass correlation coefficient of 0.035 for this analysis population, suggesting that any effect on inference is likely to be modest.

### Systematic review of EBOV-specific antibody responses after Ervebo vaccination

#### Search strategy

To identify studies reporting Ebola virus-specific antibody responses after Ervebo vaccination, we searched for relevant articles using an existing living evidence map of Ebola Virus (EBOV) studies, available at https://livingevidence.org.au/research-initiatives/about-feeva/evidence-maps/ (version 2.0, search date 1 August 2025, accessed: 2 September 2026)^11^. Specifically, we filtered on: Population = human, Intervention = vaccination, Outcomes = [Assay-Antibody binding OR Assay-Antibody neutralizing]. In addition, on 8 September 2026, we performed an updated search of the literature for studies appearing since the last update of the evidence map (on 1 August 2025). Thus, we searched PubMed and Embase from 1 August 2025 until 8 September 2026 (using the same search terms used in generating the evidence map^11,12^). The full search terms are reported in the Supplementary Methods. Studies from the evidence map and the updated search of the literature were exported to Endnote and duplicates were removed. Remaining entries were uploaded into Covidence for screening (Figure S1).

#### Inclusion and exclusion criteria and screening

The identified articles from the above searches were next screened for eligibility for inclusion. Studies were included if they reported either binding or neutralising antibody responses to EBOV (any strain) for at least one timepoint between day 7 and 14 after Ervebo/rVSV-EBOV vaccination AND for at least one timepoint 21 or more days post vaccination. We included studies of healthy adults and children (including known close contacts of Ebola virus cases) and vaccination with any vaccine dosage. Included studies were not required to include an unvaccinated or other control/comparator group. We excluded studies exclusively reporting on (i) EVD survivors, (ii) immunogenicity outcomes for other vaccines against EVD, (iii) antibody responses to VSV or other non-EBOV targets, or (iv) sero-response or sero-positivity as a binary variable. Case reports and systematic reviews were also excluded.

Screening occurred in two stages: title/abstract and full text. At both stages studies were screened independently by two reviewers (from a team of four reviewers: EE, DSK, SRK, KME). Conflicts arising during either stage of screening were resolved by discussion and consensus amongst the reviewers involved in screening.

#### Data extraction

Once eligible articles were identified, relevant data was extracted by one author and verified by another (from a team of four data extractors: AM, SRK, EE, KME). The extracted data included information about the publication (first and last author, year of publication, journal, DOI), the trial and cohort (trial number, country of trial, mean or median age, percentage female, number of vaccinations, vaccine dosage), and outcome details (assay, limit of detection of the assay, binding or neutralising response, target EBOV strain, unit of the antibody response, number of samples, days post vaccination, and antibody response with 95% confidence interval).

Conflicts were resolved by discussion, consensus, and consultation with a third reviewer where required (DSK or ES). Where data was extracted from graphs, this was done independently by two authors (from the team listed above) using WebPlotDigitizer (version 5)^10^. The geometric mean of the two extracted values was used in the analysis.

#### Risk of bias

All studies included in the analysis were assessed for risk of bias by two independent assessors (EE, DSK) using an internally developed tool based on the risk of bias domains of the Cochrane ROBINS-I tool (Supplementary Methods)^13^. We adapted the ROBINS-I tool to align with the intended aims of our systematic review and meta-analysis. Specifically, the tool was modified to assess the risk of bias in studies evaluating longitudinal changes in antibody levels within vaccinated cohorts, rather than comparison of an intervention to a comparator, for which the ROBINS-I tool was originally designed. Conflicts in risk of bias assessment were resolved by discussion and consensus between assessors.

### Survival analysis to estimate timing of vaccine efficacy

To determine whether there is evidence from the cluster-randomised controlled ring vaccination trial that vaccine protection begins earlier than 10 days post-vaccination, we used a Cox’s proportional hazard survival analysis approach (with penalised partial likelihoods – we refer to this as “the likelihood” and “Cox model” hereafter – using Firth’s method for correcting for small or no events in the vaccinated arm). Specifically, we fitted the extracted data using a Cox model and assessed the evidence that the proportional hazard assumption is violated using a Schoenfeld residual plot (Figure S2). We then fitted a time-varying Cox model with time-varying hazard ratio given by the function,

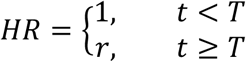

where *t* is time and *r* is the hazard ratio (HR) after a given breakpoint in time (*T*). This was fit sequentially for different values of *T* (between day 0 and day 20 – the duration of the study before the delayed arm received the vaccine). Using the likelihood of the fitted models we created a profile likelihood plot for *T*, and calculated the proportion of the area under the profile likelihood curve that includes *T* < 10. The cox model was fitted with *coxphf* function from *coxphf* package in R^14^.

Finally, using the raw case numbers that occurred in each arm from given times post randomisation in the trial, we estimated the relative risk (and accompanying 95% CI using the score-based method for relative risk with the *riskscoreci* function from the *PropCIs* package in R)^15^. We then reported the vaccine efficacy (VE) over these time intervals as VE = 1 -relative risk.

### Meta-analysis of EBOV-specific antibody responses after Ervebo vaccination

We performed two meta-regression analyses of antibody data following vaccination. Firstly, we estimated the fold-change in antibody levels between day 7 or 14 and day 28 post-vaccination across all studies where antibody levels for these time points were available. Secondly, we estimated antibody kinetics over time from all the available antibody data and timepoints included in our analysis.

#### Day 28 meta-analysis

To determine the pooled fold-change from early time points to day 28 across the included studies, we performed a multi-level random effects meta-regression using the *rma.mv* function from the *metafor* package (version 4.8-0) in R^16^. This was done separately for the fold-change from day 7 to 28 and day 14 to 28, with fold-changes calculated from the reported antibody levels. Sampling variances were calculated from reported standard errors or derived from reported 95% confidence intervals (CIs) or interquartile ranges. Where the reported antibody level was at or below either the assay limit of detection or the baseline reported antibody level, the sampling variance was assumed to be the highest variance reported for any timepoint from the same cohort (which we assumed provides a conservative estimate of the variance at these points). This alleviates the artificial reduction in sampling variances that occurs when many observations are at or below the limit of detection. The meta-regression was inverse variance weighted, with an outcome of log_10_(Ab fold-change), random intercepts for study and assay within each study (where there were multiple assays reported within a study), and a fixed effect for antibody response type (binding vs neutralising).

#### Kinetics of antibodies over time

To predict antibody levels to EBOV over time, we fitted a piecewise linear mixed effects model (using the *lmer* function in the *lme4* R package, version 1.1-37)^17^ to the extracted EBOV antibody data (log_10_-transformed) with separate early and late linear components, and inverse variance weighting of the data. This piecewise linear model allows for an initial increase in the antibody responses (data on days 7 to 14 post vaccination) and subsequent waning of the response (data from day 21 post vaccination onwards). The model contains random effects on the intercepts for both the early and late segments and on the antibody slope for the early segment (by study and assay within each study where there are data from multiple assays in a single study). The model also contains fixed effects for antibody response type (binding vs neutralising) and whether the vaccine dose is in the target dose range (i.e. 2 × 10^7^ – 1 × 10^8^ plaque-forming units (pfu)). Contrast analyses were used to estimate antibody levels at different times relative to day 7, day 10 or day 14 (using the *emmeans* function in *emmeans* R package, version 1.11.2-8)^18^. Confidence intervals were calculated from the standard errors returned by *emmeans*.

Finally, to predict BDBV antibody levels at different times after Ervebo vaccination based on the EBOV data over time and the previously reported 2.8-fold drop in binding to / neutralisation of BDBV (95% predictive interval, PI: 2.6 – 3.0)^4^, we applied this fold-drop to the estimated EBOV antibody levels over time in the above piecewise linear model. We computed confidence bands on this predicted antibody level by combining the variances in the estimates for the EBOV antibody levels over time and the uncertainty in the BDBV fold-drop estimate (assuming independence of these uncertainties).

For the meta-analysis and all statistical analyses, a significance level of 0.05 was used, i.e. p-value <0.05. All confidence and prediction intervals are 95% confidence and prediction intervals, and all statistical analyses were performed using R (version 4.5.1)^19^.

## Results

### Timing of Ervebo protection against EVD when administered in a ring-vaccination trial

The pivotal (and only) randomised clinical efficacy trial of Ervebo reported a pre-specified outcome of vaccine efficacy from day 10 post vaccination. From day 10 there were no cases in the vaccinated arm and so the reported efficacy was 100% (with reported 95% confidence interval, CI: 68.9 – 100.0)^8^. However, it is possible that protection began earlier than day 10, since the survival curves appeared to diverge from day 7 post vaccination. Thus, to explore whether vaccine efficacy may have been provided prior to day 10, we extracted the original case data from this trial (reproduced in Figure 1A) and assessed vaccine efficacy over time since Ervebo vaccination using a Cox model. Given clear evidence that the vaccine effect was not constant with time (Figure S2), we compared models in which the vaccine effect began on each day between day 0 and 20. The data provided strong support for protection beginning before day 10 (with 96% of the area under the profile likelihood corresponding to protection onset times before day 10, Figure S3). Further, although vaccine efficacy was not 100% (as is the case for vaccine efficacy after day 10), vaccine efficacy is similarly high from 7 days after vaccination where vaccine efficacy was 90.3% (95% CI: 42.4 - 98.4) (Figure 1B). Together, these results suggest that Ervebo provided protection prior to day 10 following vaccination in the pivotal ring-vaccination trial.

**Figure 1.**
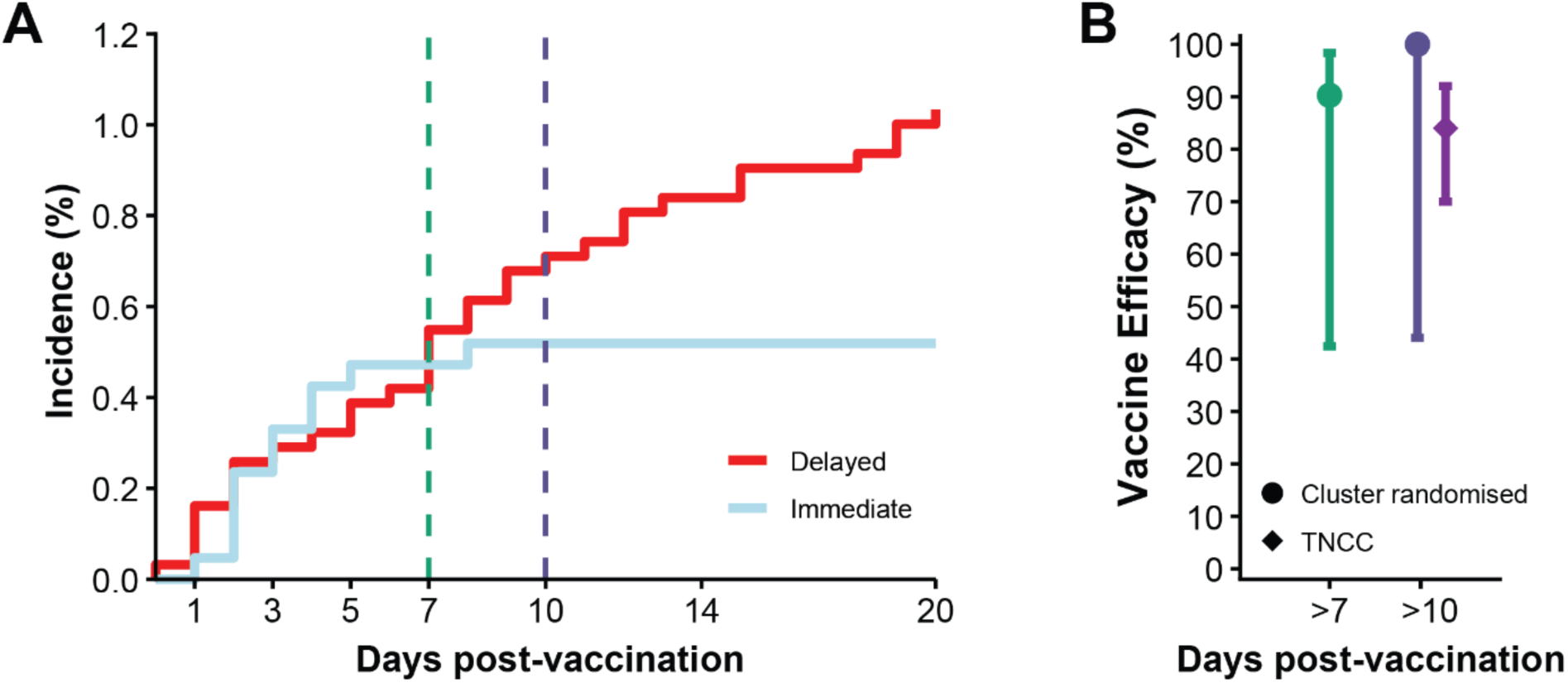
Estimating Ervebo protection from Ebola virus disease at different times after vaccination. (A) Survival data from the original ring vaccination trial was extracted for analysis (from Henao-Restrep et al. (2017)^8^). This compared infection in those vaccinated immediately (blue line) with those eligible for delayed vaccination at day 21 (red line). (B) Estimates of vaccine efficacy after specific days post-vaccination (estimates and 95% CIs indicated by circles and lines, respectively). The vaccine effectiveness from day 10 estimated in a TNCC study^20^ is also shown (purple diamond and 95% CI line). Colours (green and purple) indicate different days from which vaccine efficacy was estimated. Abbreviations: TNCC test-negative case-control study.

### Systematic review of antibody responses to EBOV after Ervebo vaccination

The analysis above suggests that early vaccine protection against Ebola virus disease could be observed on or before day 10 after Ervebo vaccination. If we assume that EBOV-specific antibody levels are associated with protection, then the antibody levels on the day that protection is conferred may reflect a putative ‘protective threshold’ sufficient to provide protection^9^ (Figure 2A,B).

**Figure 2.**
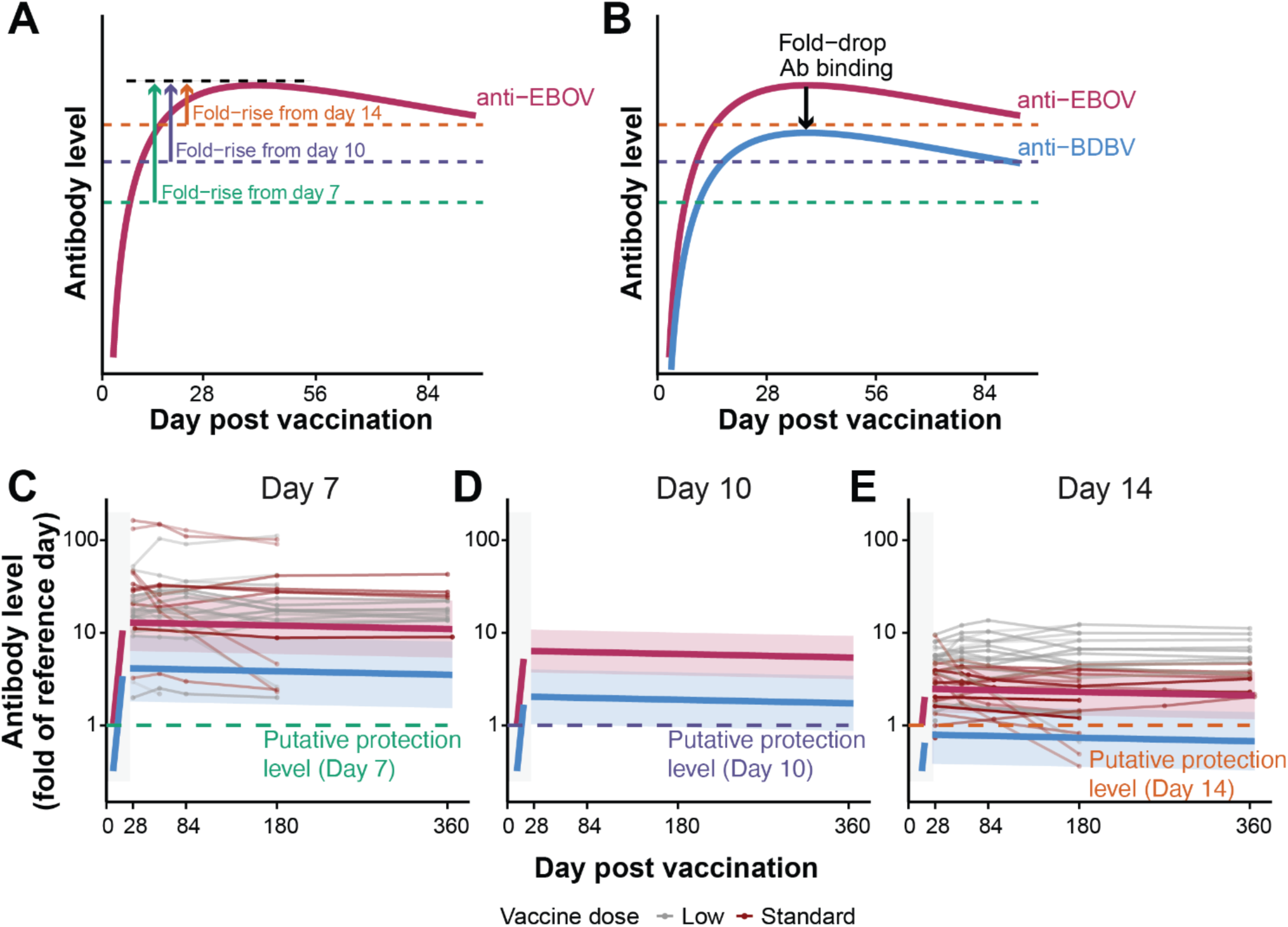
Predicting antibody levels to EBOV and BDBV over time. (A) Schematic showing fold rise in EBOV-specific antibodies from days 7, 10, and 14 post vaccination to peak at about day 28. (B) Schematic indicating a 2.8-fold drop in antibody recognition of BDBV compared to EBOV. (C-E) Predicted antibody levels to EBOV (maroon) and BDBV (blue) over time. Extracted data is shown as thin lines for individual studies (in maroon for standard doses (2 x 10^7^ – 1 x 10^8^ pfu) and grey for doses below 2 x 10^7^ pfu). Opacity of dots and lines reflects inverse variance of the antibody level. Solid maroon line and shaded area (95% CI) is shown for Ebola. Solid blue line and shaded region (95% CI) is shown for Bundibugyo. Panels (C), (D) and (E) consider antibody levels relative to day 7, 10, and 14, respectively.

We performed a systematic search of EBOV-specific antibody responses early after Ervebo vaccination. We focused on studies that reported antibody levels between day 7 and 14 post-vaccination, as well as at later times (>day 21). We identified 17 eligible publications (Figure S1). The studies used different vaccine doses (from 3 × 10^3^ to 1 × 10^8^ plaque-forming units (pfu)) and measured antibodies over different timeframes (up to 1 year post vaccination) (Table S1)^21–30^. Of these 17 studies, after excluding papers that reported the same or overlapping data with other included papers, 10 studies contained unique data and were eligible for inclusion in our analysis (Tables S1 and S2, Figure S1).

We extracted data from the 10 eligible studies for analysis^21–30^. Of these, six studies reported binding antibody responses^21–26^ and four reported both binding and neutralising antibody responses^27–30^. In our risk of bias assessment 3 of 10 studies were assessed to have a high or critical risk of bias (Figure S4). None of the included studies reported antibody levels at day 10 post vaccination (Table S1). However, one study reported responses at day 7^23^, five at day 14^24–28^, and four studies reported antibody responses on both day 7 and 14^21,22,29,30^. Nine studies reported EBOV-specific antibodies at day 28, and one study reported antibody responses at day 30^23^. Nine studies reported antibody levels to six months (180 days) post-vaccination^21–24,26–30^, and four studies reported antibody levels to one year (360 or 365 days)^23,26,28,29^ (Table S1).

### Binding and neutralising antibody responses to EBOV at peak compared to early after Ervebo vaccination

Having identified studies reporting early EBOV-specific antibody levels, we wanted to analyse how these compared to peak or later levels. Since all studies reported either a day 28 or day 30 level (which we refer to collectively as ‘day 28 level’), we compared how much higher the day 28 level was compared to the reported day 7 or day 14 EBOV-specific antibody level in the same study (Figures S5 and S6). Comparing across all studies, we found that the day 28 antibodies were on average 17.49-fold (95% CI: 5.33 – 57.40-fold) and 11.01-fold (95% CI: 2.96 – 40.95-fold) higher than the reported day 7 binding and neutralising antibody levels, respectively. Limiting our analysis to studies that used an Ervebo dose between 2 × 10^7^ – 1 × 10^8^ pfu (i.e. between the dose used in the ring-vaccination trial (2 × 10^7^pfu) and the dose approved by the European Medicine Agency (≥7.2 × 10^7^ pfu)^31^) led to similar conclusions (Table S3).

Comparing EBOV-specific day 28 antibody levels with those at day 14, we found that EBOV-specific binding antibody levels at day 28 were on average 2.55-fold higher than at day 14 (95% CI: 1.60 – 4.07-fold), and neutralising antibody levels on average 2.14-fold higher (95% CI: 1.17 – 3.92-fold). Again, when looking only at doses between 2 × 10^7^ – 1 × 10^8^ pfu, the result was similar (Table S3).

An important caveat to this analysis is that none of the five studies that reported antibody levels at day 7 post vaccination showed an increase in EBOV-specific binding or neutralising antibody levels compared to the levels reported at baseline. This might be because antibody levels had not increased, or because they were still below the level of background binding or below the limit of detection of the assay. As a result, differences between day 7 and day 28 antibodies may be even greater than shown in our analysis (because true day 7 levels may have been lower than measured).

### Predicting antibody levels to EBOV over time compared to a putative protective threshold

Analysis of the survival data from the ring vaccination trial suggested that protection from Ebola virus disease was achieved by day 10 post vaccination. Therefore, EBOV-specific antibody levels around this time might represent a putative ‘protective’ threshold level.

However, our systematic review did not identify data on antibody levels 10 days after Ervebo vaccination. Therefore, we used a piecewise linear mixed-effects model to analyse EBOV-specific antibody responses early (days 7-14) and late (days 28-365) after vaccination (Figure S7). This allowed us to interpolate EBOV-specific antibody levels on day 10. We then compared the long-term levels of EBOV-specific antibodies with levels at either day 7, day 10 (interpolated from the data), or day 14 (Figure 2C-E). This suggests that long-term EBOV-specific antibody levels are maintained well above all these potential protective thresholds for at least a year (Figure 2C-E, maroon-coloured lines).

### Predicting Ervebo protection from Bundibugyo virus disease

To predict Ervebo protection from Bundibugyo virus disease, we need to understand how BDBV-specific antibody levels after vaccination relate to the putative protective antibody levels determined for Ebola (Figure 2B). We have previously estimated that cross-reactive antibodies to Bundibugyo virus (BDBV) (or the BDBV GP) are 2.8-fold (95% prediction interval, PI: 2.6 – 3.0) lower than to EBOV^4^ (based on analysis of data from day 21 – 660 after Ervebo vaccination). We can use this drop in antibody recognition to predict how BDBV-specific antibody levels compare to the putative protective EBOV-specific antibody levels at day 7, 10 or 14 post vaccination (Figure 2, blue coloured lines). If the antibody levels to EBOV at day 7, 10 or 14 do indeed represent a protective threshold against Ebola virus disease, and assuming the same protective threshold also applies for Bundibugyo virus disease, we can then predict whether BDBV-specific antibody levels are in the protective range. We find that if the EBOV-specific antibody level at day 7 is protective, then antibodies to BDBV are predicted to rise above this putative protective level by day 28 and remain above this level durably (Figure 2C). If the EBOV-specific antibody level at day 10 is protective, we predict that the BDBV antibody level probably rises above this putative protective level and remains high. However, the 95% CIs include the protective threshold (Figure 2D). Finally, if the EBOV-specific antibody level at day 14 were the protective threshold, then it is unlikely the BDBV antibody levels ever rise above this putative protective threshold (Figure 2E).

## Discussion

The goal of this study was to place available evidence on immune cross-reactivity to BDBV after Ervebo vaccination in the context of the established protection of the Ervebo vaccine against Ebola virus disease. This study does not aim to assert whether this evidence is sufficient to support the use of Ervebo to prevent Bundibugyo virus infection or death. Instead, it aims to articulate the totality of existing evidence on cross-reactivity and clearly articulate the strengths, limitations, and assumptions needed to use this data to infer potential vaccine protection against Bundibugyo virus disease. Whether this evidence is deemed sufficient for decision-making is dependent on the outbreak context and the specific risks and benefits of a decision to deploy or not deploy the vaccine.

Inferring vaccine protection from immunogenicity data can be challenging even for a well-studied infection. Attempting to do this based on a combination of protection against a different strain of the virus and cross-reactivity of antibodies to the two virus strains adds an additional level of indirectness to any inference. In the absence of clinical data on protection against Bundibugyo virus disease after Ervebo vaccination, this study analyses data on immune responses to EBOV and protection from Ebola virus disease after Ervebo vaccination and then incorporates the degree of immune cross-reactivity to BDBV to infer protection against Bundibugyo virus disease. Inferring this protection relies on a number of important assumptions that need to be clearly stated. Firstly, a critical assumption is that EBOV-specific antibody levels after Ervebo vaccination are predictive of clinical protection from Ebola virus disease. There is no direct evidence that EBOV-specific antibodies predict protection from Ebola virus disease after Ervebo vaccination in humans. However, studies of monoclonal antibody therapy of Ebola virus-infected individuals demonstrate that high-dose antibodies can protect from death (although convalescent serum was not shown to be effective)^32,33^. Studies have also shown an association between binding antibody levels and protection from death after EBOV challenge of non-human primates (NHPs) after both rVSV-EBOV-GP and Zabdeno/Mvabea vaccination, supporting a role for antibodies in predicting protection in NHPs^34–38^.

Inferring Ervebo protection against Bundibugyo virus disease requires a further assumption that BDBV-specific antibodies are also predictive of protection from Bundibugyo virus disease. In addition, it relies on the assumption that the relationship between virus-specific antibodies and protection from disease for Bundibugyo virus is the same as for EBOV. There are currently no studies demonstrating an association between BDBV-specific immune responses and protection from infection in humans. However, monoclonal antibodies have been shown to be effective in preventing death in studies of post-exposure therapy in both ferret and non-human primate models of Bundibugyo infection^39–41^. There are also several studies comparing serological responses to BDBV after vaccination and protection from infection and death in non-human primates, although at present there is no definitive evidence of an association^42,43^.

A strength of this study is the aggregation of all relevant data on the kinetics of immune responses after Ervebo vaccination through a systematic review and meta-analysis. This is coupled with a previous systematic study of antibody cross-reactivity after Ervebo vaccination^4^. However, the data aggregation approach we have used has its own limitations because the analysis involves aggregation of data from quite disparate clinical study types and *in vitro* immune assays. The primary studies from which the data were extracted were also often not designed to address the specific questions raised in our study. Immune responses were typically measured either using binding assays or *in vitro* neutralisation assays. *In vitro* neutralisation assays may more directly assess antiviral function, but were only used in the minority of studies of both antibody kinetics and cross-reactivity (see Stadler et al. (2026)^4^ and Table S1). However, the conclusions regarding antibody levels at different times were not significantly different between binding and neutralising assays. Finally, we have assessed clinical study design using a custom-made risk of bias assessment tool with domains adapted from the ROBINS-I tool (Supplementary Methods), which is not an established tool for assessing risk of bias. However, we are not aware of an established risk of bias tool for the question considered in this study. For these reasons, the risk of bias assessment provided here should be interpreted with caution.

The analysis above primarily considers long-term protection against Ebola and Bundibugyo virus disease (post day 28). However, planned trials of vaccination for Bundibugyo rely on ring vaccination of contacts and thus are dependent on early vaccine effectiveness. What would a 2.8-fold drop in antibody recognition mean for the effectiveness of the vaccine in these trials? If we assume the kinetics of cross-reactive antibodies to Bundibugyo are the same as those to Ebola but at 2.8-fold lower levels, we can estimate the delay until any given antibody protective threshold is reached. Using the kinetics observed in our analysis (Figure S7), we estimate that for a 2.8-fold drop in specific antibody levels, the delay would be around 4.4 days. In a clinical trial of Ervebo protection from Bundibugyo virus disease, if the primary outcome is protection from Bundibugyo virus disease after day 10, then delay in the onset of protection may reduce the observed vaccine effectiveness (depending on when protection is initially achieved). However, this prediction relies on a number of assumptions, including that cross-reactivity against BDBV very early after vaccination is the same as that observed later. At present there is no data to address this question. However, considering the possibility of a delay in onset of protection is likely important in powering and analysing any proposed ring vaccination trials of Ervebo against Bundibugyo virus disease.

Some questions that arise from the analysis provided here include whether we would predict similar results in children, and whether the predicted protection may be improved by use of a two-dose schedule. In our systematic review of EBOV-specific responses after Ervebo vaccination, two papers included disaggregated data on children as well as adults^21,26^. From these two studies we observe that children have similar, or potentially higher, antibody levels at day 28 post-vaccination (GMT of 1503 for adults and 1620 for children from Agnandji et al. (2017)^21^, GMT of 1218 for adults and 1688 for children from Kieh et al. (2022)^26^). Further, we identified two studies that reported data for antibody levels after two doses of Ervebo compared with only 1 dose^26,30^. These two studies indicated that EBOV-specific antibody levels may transiently rise as much as 3.8-fold after receiving a second dose (3.8-fold^26^ and 1.6 to 3.8-fold^30^). However, by 6 months post vaccination antibodies had returned to levels similar to those seen after a single dose. These studies suggest that boosting with Ervebo may increase antibody responses (at least temporarily)^26,30^, although our meta-analysis of cross-reactivity suggests there may also be less cross reactivity with BDBV after boosting (trend only, not significant, based on a single study)^4^.

Our analysis of the available data on immune responses to Bundibugyo and Ebola highlights a number of gaps, which future studies could address. Firstly, the lack of detailed serological data early after Ervebo vaccination presents a challenge in understanding what might be the protective antibody levels around this time (e.g. at day 10). This is compounded by sensitivity and/or specificity limits of the assays for detecting responses early after vaccination. Therefore, at present there is a lack of quantitative measurement of serological responses before day 14 post-vaccination. Further data on immune responses and cross-reactivity after boosting and in children would also be helpful.

The primary aim of this work has been to highlight the required assumptions and limitations of the available immunological data for determining whether Ervebo will likely be protective against Bundibugyo virus disease, in order to better support decision-makers. Depending on the validity of the many assumptions invoked, our analysis of the immunological data may leave some readers optimistic on the potential benefit of Ervebo as a pre-exposure prophylaxis against BVD. Further immunological studies may strengthen some of the assumptions here. However, randomised trials to demonstrate clinical protection are urgently needed.

## Data Availability

All data and code are available upon request to the authors and will be made publicly available upon publication.

## Acknowledgements

The authors wish to thank Steve McDonald for running the updated Ebola search and Arnaud Marchant for helpful comments on the manuscript. This work is funded by the National Health and Medical Research Council (NHMRC, Australia) Investigator Grants 2034282 (to ES), 2026360 (to DC), 2034108 (to MPD), 2033318 (to DSK). This research has been supported by the Australian Commonwealth Government through an Australian Government Research Training Program (RTP) Scholarship [DOI: https://doi.org/10.82133/C42F-K220] (to KME).

## Ethics statement

This work was approved under the UNSW Sydney Human Research Ethics Committee (approval HC200242).

## Contributions

Conceptualization: MPD, DSK, ES

Data curation: KME, EE, SRK, AM, AR, ES, DSK

Risk of bias: EE, DSK

Formal analysis: KME, AR, ES

Funding acquisition: DC, ES, MPD, DSK

Methodology: KME, AR, MTB, TES, DC, TT, ES, DSK

Project administration: BAB

Supervision: DC, ES, MPD, DSK

Writing – original draft: DC, ES, MPD, DSK

Writing – reviewing & editing: All authors

