## Supplementary Material for "Predicting the efficacy of Ervebo against Bundibugyo virus disease"

### Table of contents

**Supplementary Methods** **2**

Full search terms for updated EBOV search 2
Risk of bias assessment tool for immunogenicity studies 3

**Supplementary Figures** **5**

Figure S1. PRISMA flowchart for the systematic search of antibody responses to Ebola virus after Ervebo vaccination 6
Figure S2. Schoenfeld residual plot 7
Figure S3. Profile likelihood 8
Figure S4. Risk of bias assessment outcomes 9
Figure S5. Forest plot of fold-change in EBOV-specific antibody responses from day 7 to day 28 10
Figure S6. Forest plot of fold-change in EBOV-specific antibody responses from day 14 to 28 12
Figure S7. Antibodies over time by study and assay fitted with a piecewise linear mixed-effects model 13

**Supplementary Tables** **14**

Table S1. Characteristics of studies and cohorts included in the analysis 14
Table S2. Studies identified in the systematic review but not included in the analysis 17
Table S3. Fold-change estimates between day 7 or 14 and day 28 using all data compared with restricting to include data on the standard doses only 18

**References** **19**

### Supplementary Methods

#### Full search terms for updated EBOV search

The evidence map used to identify studies https://livingevidence.org.au/research-initiatives/about-feeva/evidence-maps/ (version 2.0, search date 1 August 2025, accessed 2 September 2026)[1], was last updated on 1 August 2025. In order to identify more recent studies, we performed additional literature searches of the databases PubMed and Embase. We used the same search queries as those used to generate the EBOV evidence map, and these are provided in full below for each database:

**PubMed**

(Filoviridae[mh:noexp] OR Filoviridae Infections[mh:noexp] OR Ebolavirus[mh] OR Hemorrhagic Fever, Ebola[mh] OR ebola*[ti] OR EBOV[ti] OR ((ebola*[tiab] OR EBOV[tiab]) NOT medline[sb])) AND (Ebola Vaccines[mh] OR Vaccine Development[mh] OR Vaccination[mh] OR vaccin*[ti] OR (vaccin*[tiab] NOT medline[sb]) OR Disease Models, Animal[mh] OR Enzyme-Linked Immunosorbent Assay[mh] OR Antibodies, Neutralizing[mh] OR Antibodies, Monoclonal[mh] OR Neutralization Tests[mh] OR Immunity[mh] OR ELISA*[tiab] OR immuno*[tiab] OR immune*[tiab] OR neutrali*[tiab] OR monoclonal[tiab] OR mAbs[tiab] OR antiviral*[tiab] OR assay*[tiab] OR T-cell*[tiab] OR "animal model"[tiab:~5] OR "animal models"[tiab:~5] OR non-human*[tiab] OR nonhuman*[tiab] OR primate*[tiab] OR chimpanzee*[tiab] OR macaque*[tiab] OR mice[tiab] OR mouse[tiab] OR hamster*[tiab] OR ferret*[tiab] OR murine[tiab] OR "guinea pig*"[tiab] OR "animal challenge"[tiab] OR "lethal challenge"[tiab] OR "human challenge"[tiab] OR "infection challenge"[tiab] OR "natural history"[tiab]) NOT (Review[pt] OR News[pt] OR Editorial[pt])

**Embase Classic and Embase**

| # | Search Statement |
|---|---|
| 1 | exp *Ebolavirus/ or *Ebola Hemorrhagic Fever/ or (ebola* or EBOV).ti,kf |
| 2 | Ebola Vaccine/ or Vaccine Development/ or Vaccination/ or vaccin*.ti,ab,kf. |
| 3 | Disease Model/ or exp Enzyme Linked Immunosorbent Assay/ or exp Neutralizing Antibody/ or exp Neutralization Test/ or (ELISA* or immuno* or immune* or neutrali* or monoclonal or mAbs or antiviral* or assay* or T-cell* or (animal adj5 model*) or non-human* or nonhuman* or primate* or chimpanzee* or macaque* or mice or mouse or hamster* or ferret* or murine or guinea pig* or (animal or lethal or human or infection adj1 challenge) or natural history).ti,ab. |
| 4 | 1 and (2 or 3) |
| 5 | exp Review/ or (Editorial or Note or Conference Abstract or Clinical Trial).pt. |
| 6 | 4 not 5 |
| 7 | limit 6 to (english language and "remove medline records") |

**Risk of bias assessment tool for immunogenicity studies**

Due to the lack of a tool designed for assessing risk of bias for studies of longitudinal changes in immunogenicity, we developed a risk of bias (RoB) tool based on the risk of bias domains of the Cochrane ROBINS-I tool[2].
The modified tool maintains the domain structure of ROBINS-I but is simplified and adapted to be appropriate for our research question (i.e. longitudinal changes in antibody levels rather than an effect from an intervention compared to a control group).

The domains and accompanying considerations are described below:

***Adapted RoB tool for assessing included studies***
For each domain:

- Yes = **LOW RoB.**
- Unclear or No, but unlikely to have substantive impact on results = **MODERATE RoB.**
- No, and likely to have substantive impact on results = **HIGH RoB.**
- No, and likely to have major impact on results = **CRITICAL RoB.**

**D1: Bias due to confounding**

- Were all data collected from one group of participants, recruited contemporaneously, with a single set of inclusion criteria and outcome measurement timepoints and definitions?

**D2: Bias in classification of interventions**

- Did all participants receive the intervention to which they were allocated, OR are they analysed as if they received the intervention to which they were allocated?

**D3: Bias in selection of participants into the study (or into the analysis)**

- Selection of participants is not influenced by any characteristic that might impact on the findings (particularly outcome or confounding variables)?

**D4: Bias due to missing data**

- Outcome data is available for all outcomes for all participants who were selected to participate and at all included timepoints? **OR**

  Patterns in missing data are similar across groups and timepoints and are likely to be random **AND**

- Methods for imputing missing data are appropriate?

**D5: Bias in measurement of the outcome**

- Outcome measurement is blinded (participants AND outcome assessors) **OR** Outcome measures are objective, and all reported outcomes are measured in the same manner and at the same timepoint in all participants

**D6: Bias in selection of the reported result**

- There is a protocol and the outcomes reported and analysis methods used are as specified in the protocol?

**Overall RoB assessment**

- Overall result is equal to highest domain RoB rating.
- If the there are two or more ratings at the highest level, round up to next level (e.g. three Moderate domain ratings, becomes a High overall rating)

### Supplementary Figures

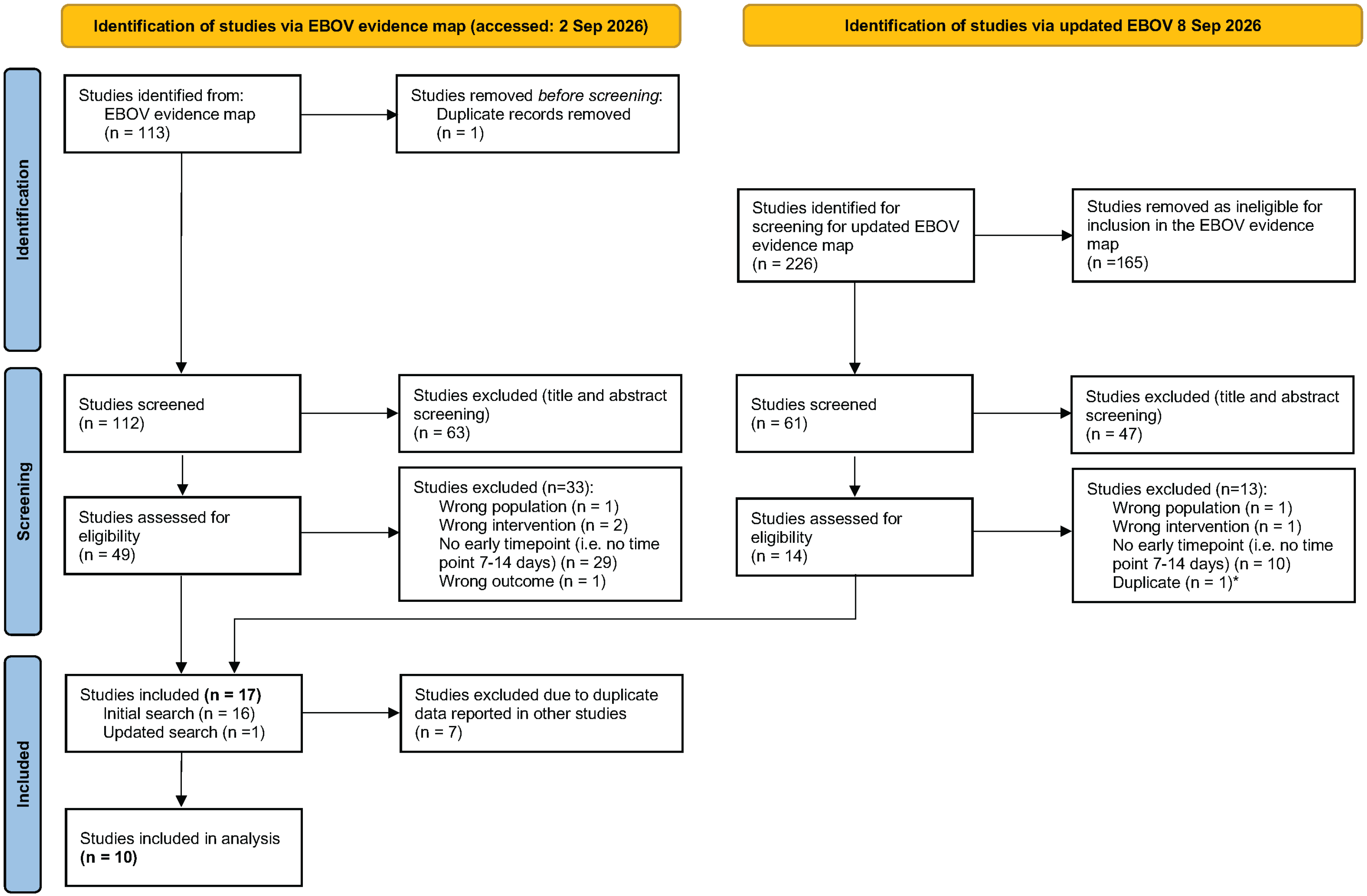

**Figure S1. PRISMA flowchart for the systematic search of antibody responses to Ebola virus after Ervebo vaccination**

Left hand side: Searches of the EBOV evidence map (https://livingevidence.org.au/research-initiatives/about-feeva/evidence-maps/), which has a search date of 1 August 2025 and was accessed on 2 September 2026. Right hand side: Updated search using the evidence map search terms, performed on 8 September 2026
*When screening articles in the updated search (right), one study was found to meet the inclusion criteria at full-text review, but was subsequently identified as an exact duplicate of a study already identified and included from the search of the evidence map (left).

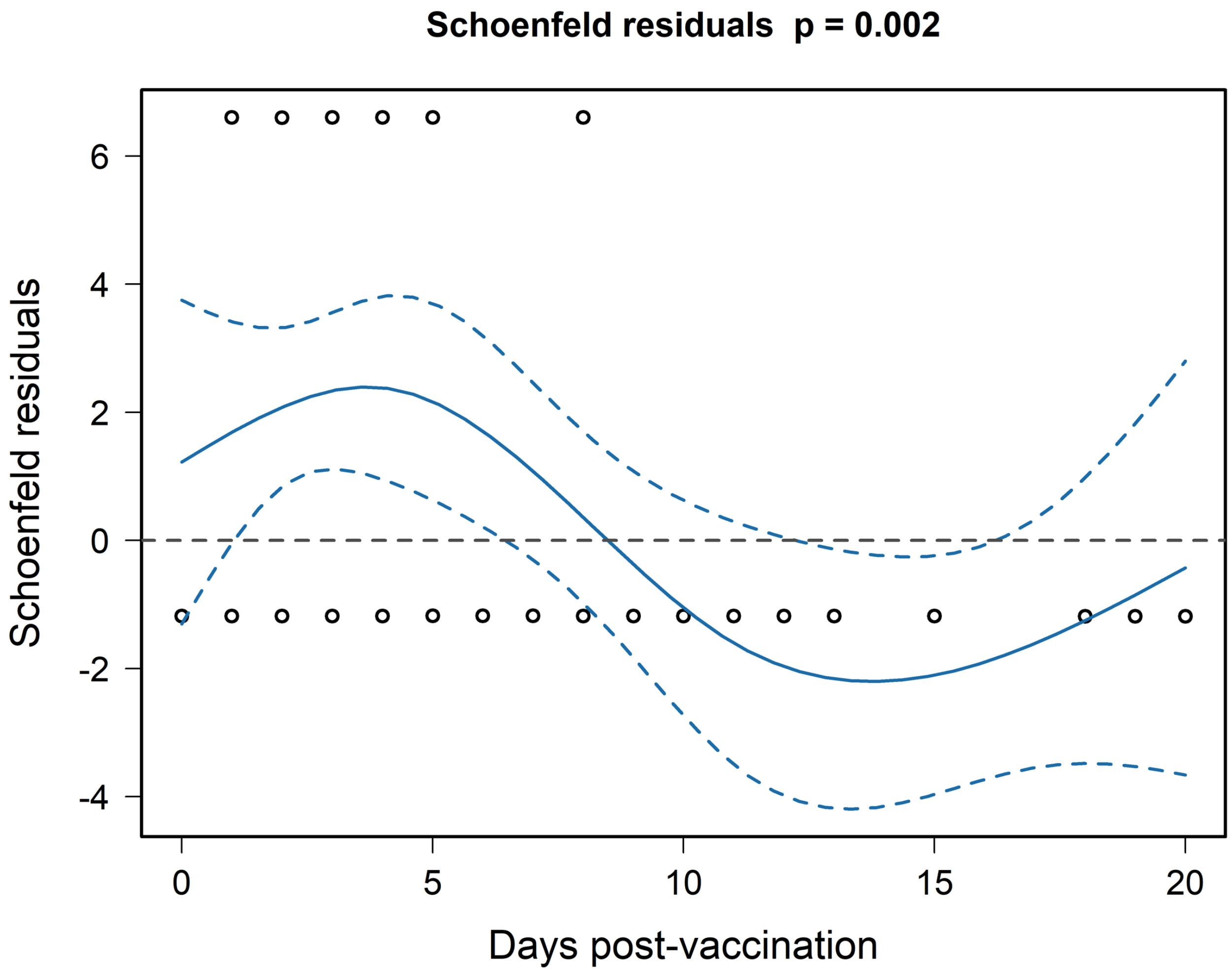

**Figure S2. Schoenfeld residual plot**

We first fitted a penalised Cox proportional hazard model (Firth's correction – to account for the small number of events in the vaccine arm) to the data in Figure 1 of the main text. This Schoenfeld residual plot shows strong evidence that the proportional hazard assumption is violated, highlighting evidence that the hazard ratio is not constant with time.

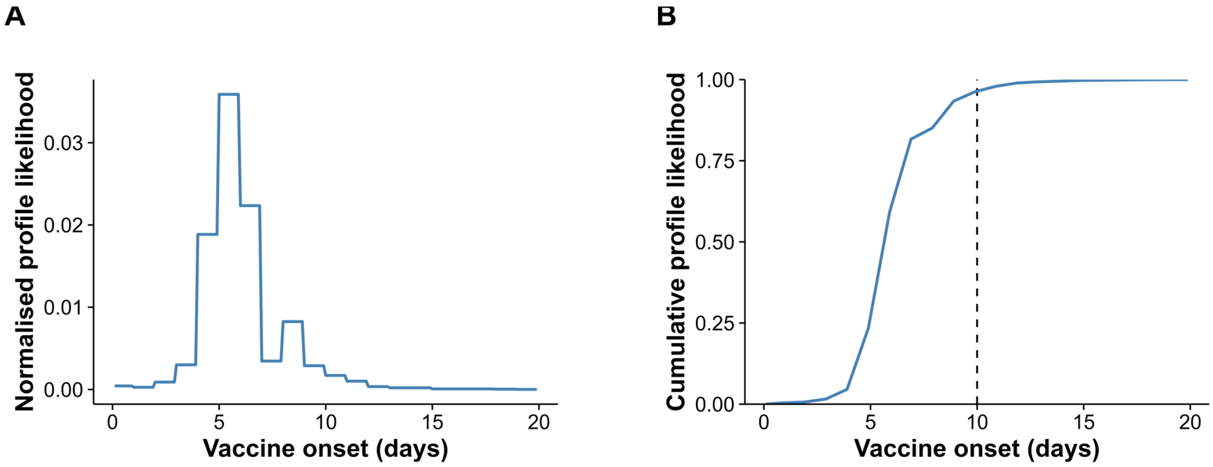

**Figure S3. Profile likelihood**

(A) Here we show the penalised partial profile likelihood from fitting a time-varying Cox model (with Firth's correction) to the data in Figure 1A of the main text. The time-varying Cox model has a hazard ratio fixed to 1 prior to a time $T$ and a constant hazard ratio (estimated by model fitting) after time $T$ (where $T$ is a parameter that describes the time at which the vaccine began to provide protection - vaccine onset day). The profile likelihood has been normalised so that the area under the curve is equal to 1. (B) We show the corresponding cumulative density of the profile likelihood. There is strong support that the time at which vaccine efficacy begins is before day 10.

| | Risk of bias domains | | | | | | |
|---|---|---|---|---|---|---|---|
| Study | D1 | D2 | D3 | D4 | D5 | D6 | Overall |
| Agnandji 2017 | + | + | + | + | + | - | - |
| Agnandji 2016 | + | + | + | + | + | + | + |
| Boum 2020 | + | + | X | X | - | + | X |
| Davis 2020 | + | + | - | X | - | + | X |
| ElSherif 2017 | + | + | + | + | + | + | + |
| Heppner 2017 | + | + | + | + | + | + | + |
| Huttner 2015 | X | + | + | - | + | + | X |
| Kennedy 2017 | + | + | + | + | + | + | + |
| Kieh 2022 | + | + | + | + | + | + | + |
| Regules 2017 | + | + | + | + | + | + | + |

**Domains**
**D1:** Bias due to confounding
**D2:** Bias in classification of interventions
**D3:** Bias in selection of participants into the study (or into the analysis)
**D4:** Bias due to missing data
**D5:** Bias in measurement of the outcome
**D6:** Bias in selection of the reported result

| **Judgement** | |
|---|---|
| + | Low risk of bias |
| - | Moderate risk of bias |
| X | Serious risk of bias |
| X | Critical risk of bias |
| ? | No information |

**Figure S4. Risk of bias assessment outcomes**

Fold−change from day 7 to day 28

| Study | Assay notes | N | dose (PFU) | Fold−change (95%CI) |
|---|---|---|---|---|
| Heppner et al. 2017 | GP ELISA IgG | 64 | 3000 | 17.02 (11.45−25.32) |
| Heppner et al. 2017 | GP ELISA IgG | 61 | 30000 | 14.91 (10.38−21.42) |
| ElSherif et al. 2017 | GP ELISA IgG | 10 | 1e+05 | 22.97 (6.14−85.89) |
| Agnandji et al. 2017 | Whole virion ELISA IgG | 20 | 3e+05 | 2.94 (0.56−15.60) |
| Heppner et al. 2017 | GP ELISA IgG | 64 | 3e+05 | 20.99 (15.49−28.45) |
| ElSherif et al. 2017 | GP ELISA IgG | 10 | 5e+05 | 21.79 (5.40−87.93) |
| Agnandji et al. 2017 | Whole virion ELISA IgG | 39 | 3e+06 | 1.99 (1.30−3.05) |
| ElSherif et al. 2017 | GP ELISA IgG | 10 | 3e+06 | 47.70 (20.29−112.12) |
| Heppner et al. 2017 | GP ELISA IgG | 83 | 3e+06 | 25.08 (19.11−32.91) |
| Regules et al. 2017 | GP ELISA IgG | 20 | 3e+06 | 52.00 (20.74−130.39) |
| Heppner et al. 2017 | GP ELISA IgG | 45 | 9e+06 | 16.21 (10.87−24.17) |
| Agnandji et al. 2017 | Whole virion ELISA IgG | 16 | 2e+07 | 3.24 (1.39−7.60) |
| Heppner et al. 2017 | GP ELISA IgG | 46 | 2e+07 | 29.64 (20.54−42.78) |
| Kennedy et al. 2017# | GP ELISA IgG | 491 | 2e+07 | 11.12 (9.69−12.76) |
| Regules et al. 2017 | GP ELISA IgG | 20 | 2e+07 | 163.16 (67.85−392.36) |
| Heppner et al. 2017 | GP ELISA IgG | 45 | 1e+08 | 28.28 (19.35−41.31) |
| Regules et al. 2017 | GP ELISA IgG | 20 | 1e+08 | 132.52 (46.66−376.37) |
| **Pooled estimate** | **All binding** | | | **17.49 (5.33−57.40)** |
| Heppner et al. 2017 | Pseudovirus plaque reduction | 63 | 3000 | 10.29 (5.90−17.95) |
| Heppner et al. 2017 | Pseudovirus plaque reduction | 61 | 30000 | 9.24 (5.17−16.51) |
| Heppner et al. 2017 | Pseudovirus plaque reduction | 64 | 3e+05 | 14.85 (8.87−24.89) |
| Heppner et al. 2017 | Pseudovirus plaque reduction | 83 | 3e+06 | 17.98 (12.56−25.75) |
| Regules et al. 2017 | Pseudovirus neutralisation | 20 | 3e+06 | 22.30 (7.65−65.00) |
| Heppner et al. 2017 | Pseudovirus plaque reduction | 45 | 9e+06 | 15.19 (9.14−25.26) |
| Heppner et al. 2017 | Pseudovirus plaque reduction | 46 | 2e+07 | 20.49 (12.29−34.17) |
| Regules et al. 2017 | Pseudovirus neutralisation | 20 | 2e+07 | 44.10 (15.49−125.56) |
| Heppner et al. 2017 | Pseudovirus plaque reduction | 45 | 1e+08 | 33.60 (21.30−53.01) |
| Regules et al. 2017 | Pseudovirus neutralisation | 20 | 1e+08 | 46.10 (18.92−112.34) |
| **Pooled estimate** | **All neutralisation** | | | **11.01 (2.96−40.95)** |

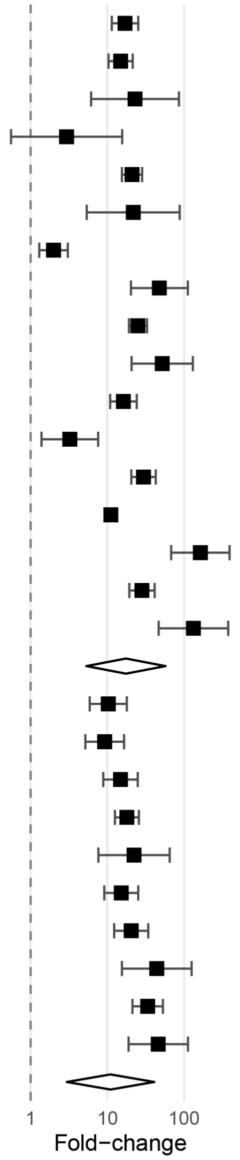

Level 2 $I^2$: 5.7%
Level 3 $I^2$: 91.6%
Test for Residual Heterogeneity: QE(df = 25) = 247.64, p < 0.0001
Test of Moderators: QM(df = 1) = 1.57, p = 0.21

**Figure S5. Forest plot of fold-change in EBOV-specific antibody responses from day 7 to day 28**

#Antibody levels for this study were reported at day 30.
Abbreviations: PFU = Plaque forming units.

### Fold−change from day 14 to day 28

| Study | Assay notes | N | dose (PFU) | Fold−change (95%CI) |
|---|---|---|---|---|
| Heppner et al. 2017 | GP ELISA IgG | 64 | 3000 | 9.53 (5.84−15.58) |
| Heppner et al. 2017 | GP ELISA IgG | 61 | 30000 | 5.74 (3.60−9.16) |
| ElSherif et al. 2017 | GP ELISA IgG | 10 | 1e+05 | 6.57 (1.76−24.58) |
| Agnandji et al. 2017 | Whole virion ELISA IgG | 20 | 3e+05 | 2.80 (0.53−14.86) |
| Heppner et al. 2017 | GP ELISA IgG | 64 | 3e+05 | 8.05 (5.32−12.18) |
| Huttner et al. 2015 | GP ELISA IgG | 51 | 3e+05 | 2.58 (1.78−3.73) |
| Huttner et al. 2015 | Whole virion ELISA IgG | 51 | 3e+05 | 1.12 (0.93−1.36) |
| ElSherif et al. 2017 | GP ELISA IgG | 10 | 5e+05 | 5.46 (1.35−22.04) |
| Agnandji et al. 2017 | Whole virion ELISA IgG | 39 | 3e+06 | 1.39 (0.85−2.29) |
| ElSherif et al. 2017 | GP ELISA IgG | 10 | 3e+06 | 7.05 (3.83−12.99) |
| Heppner et al. 2017 | GP ELISA IgG | 83 | 3e+06 | 5.52 (3.90−7.83) |
| Regules et al. 2017 | GP ELISA IgG | 20 | 3e+06 | 4.59 (2.11−9.99) |
| Heppner et al. 2017 | GP ELISA IgG | 45 | 9e+06 | 3.80 (2.25−6.44) |
| Agnandji et al. 2016 | Whole virion ELISA IgG | 27 | 1e+07 | 1.72 (1.17−2.55) |
| Huttner et al. 2015 | GP ELISA IgG | 34 | 1e+07 | 2.82 (1.76−4.51) |
| Agnandji et al. 2017 | Whole virion ELISA IgG | 16 | 2e+07 | 1.84 (0.79−4.31) |
| Boum et al. 2020 | GP ELISA IgG | 1052 | 2e+07 | 2.03 (1.88−2.18) |
| Boum et al. 2020 | Whole virion ELISA IgG | 108 | 2e+07 | 0.81 (0.61−1.09) |
| Davis et al. 2020 | GP ELISA IgG | 15 | 2e+07 | 1.85 (0.82−4.18) |
| Heppner et al. 2017 | GP ELISA IgG | 46 | 2e+07 | 3.81 (2.25−6.46) |
| Regules et al. 2017 | GP ELISA IgG | 20 | 2e+07 | 4.76 (2.36−9.58) |
| Agnandji et al. 2016 | Whole virion ELISA IgG | 12 | 5e+07 | 1.34 (0.69−2.59) |
| Huttner et al. 2015 | GP ELISA IgG | 13 | 5e+07 | 3.17 (1.69−5.91) |
| Kieh et al. 2022[a] | GP ELISA IgG | 372 | 9.4e+07 | 2.86 (2.49−3.29) |
| Kieh et al. 2022[b] | GP ELISA IgG | 396 | 9.4e+07 | 3.94 (3.42−4.55) |
| Kieh et al. 2022[c] | GP ELISA IgG | 188 | 9.4e+07 | 2.73 (2.22−3.36) |
| Kieh et al. 2022[d] | GP ELISA IgG | 191 | 9.4e+07 | 3.49 (2.87−4.26) |
| Heppner et al. 2017 | GP ELISA IgG | 45 | 1e+08 | 3.86 (2.49−5.98) |
| Regules et al. 2017 | GP ELISA IgG | 20 | 1e+08 | 4.59 (2.08−10.13) |
| **Pooled estimate** | **All binding** | | | **2.55 (1.60−4.07)** |
| Heppner et al. 2017 | Pseudovirus plaque reduction neutralisation | 63 | 3000 | 7.98 (4.57−13.91) |
| Heppner et al. 2017 | Pseudovirus plaque reduction neutralisation | 61 | 30000 | 5.70 (3.19−10.19) |
| Heppner et al. 2017 | Pseudovirus plaque reduction neutralisation | 64 | 3e+05 | 5.15 (3.29−8.06) |
| Heppner et al. 2017 | Pseudovirus plaque reduction neutralisation | 83 | 3e+06 | 5.09 (3.72−6.98) |
| Regules et al. 2017 | Pseudovirus neutralisation | 20 | 3e+06 | 5.72 (3.02−10.84) |
| Heppner et al. 2017 | Pseudovirus plaque reduction neutralisation | 45 | 9e+06 | 3.74 (2.43−5.75) |
| Boum et al. 2020 | Pseudovirus plaque reduction neutralisation | 1053 | 2e+07 | 1.61 (1.49−1.74) |
| Boum et al. 2020 | Whole virion neutralisation | 108 | 2e+07 | 0.73 (0.61−0.88) |
| Davis et al. 2020 | Whole virion neutralisation | 15 | 2e+07 | 1.00 (0.61−1.62) |
| Heppner et al. 2017 | Pseudovirus plaque reduction neutralisation | 46 | 2e+07 | 2.97 (1.78−4.96) |
| Regules et al. 2017 | Pseudovirus neutralisation | 20 | 2e+07 | 9.38 (3.30−26.71) |
| Heppner et al. 2017 | Pseudovirus plaque reduction neutralisation | 45 | 1e+08 | 3.67 (2.33−5.79) |
| Regules et al. 2017 | Pseudovirus neutralisation | 20 | 1e+08 | 3.63 (1.71−7.70) |
| **Pooled estimate** | **All neutralisation** | | | **2.14 (1.17−3.92)** |

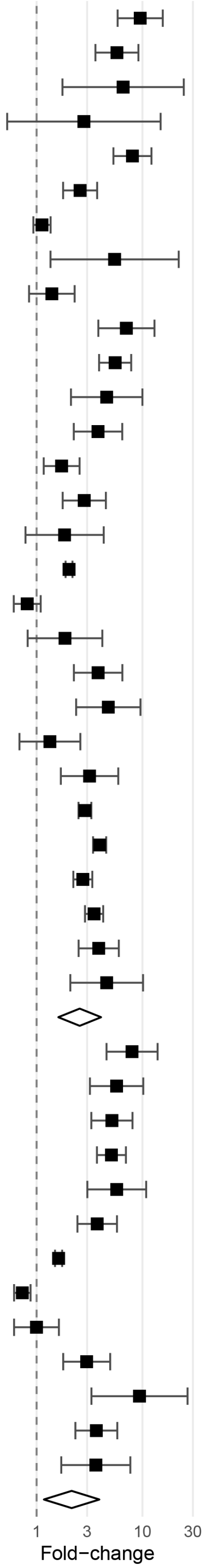

Level 2 $I^2$: 36.83%
Level 3 $I^2$: 59.68%
Test for Residual Heterogeneity: QE(df = 40) = 609.94, $p < 0.0001$
Test of Moderators: QM(df = 1) = 0.39, $p = 0.53$

**Figure S6. Forest plot of fold-change in EBOV-specific antibody responses from day 14 to 28**

[a] This cohort is adults.
[b] This cohort is children.
[c] This cohort is adults who subsequently received a 2nd dose.
[d] This cohort is children who subsequently received a 2nd dose.
Abbreviations: PFU = Plaque forming units.

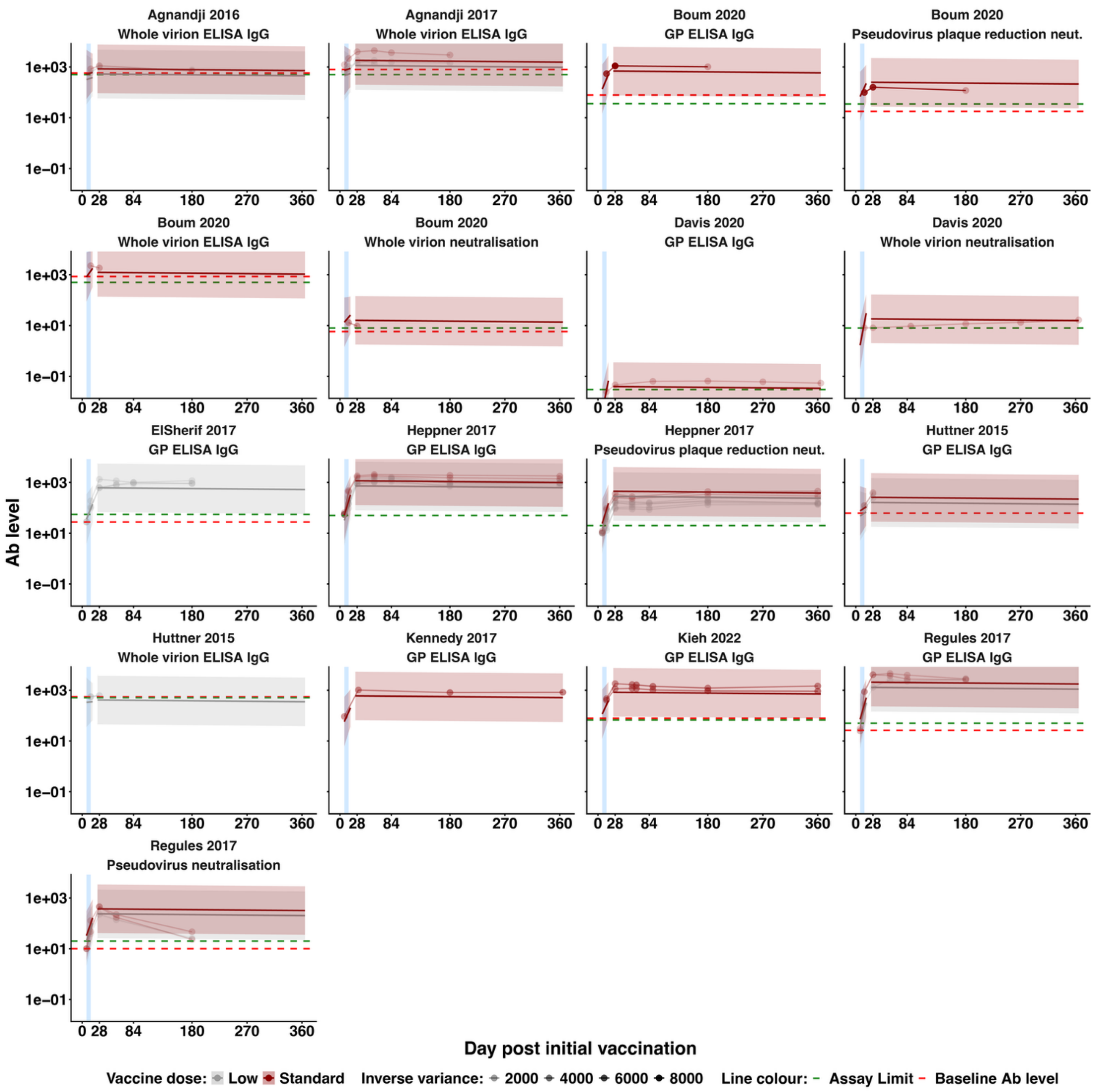

**Figure S7. Antibodies over time by study and assay fitted with a piecewise linear mixed-effects model**

Circles joined by thin lines indicate the data (extracted geometric mean antibody levels from the studies for each included cohort and assay). Opacity of the included data indicates the inverse variance of these data. Thicker solid lines indicate the fitted model for each assay and cohort from each study (fitted model lines include estimated random effects). Shaded red and grey regions indicate the 95% Confidence Intervals of the fitted model. Blue shaded regions indicate the day 7-14 time period. Red lines and points show the data and model for doses between 2 x $10^7$ and 1 x $10^8$, and the grey lines and points show data for lower doses (<2 x $10^7$). The dashed green line indicates the reported assay limit of detection from the corresponding study, and the dashed red line indicated the reported baseline antibody levels of the cohort or an accompanying naïve cohort.

### Supplementary Tables

#### Table S1. Characteristics of studies and cohorts included in the analysis

| Author (year) | Study design | Trial number | Country | Age (years, mean/ median) | Female (%) | Dose (PFU) | Sample size (range) | Assay description | Time points (days post vaccine) |
|---|---|---|---|---|---|---|---|---|---|
| Agnandji (2017)[3*] | Phase 1 randomised trial | PACTR201411000919191 | Gabon | 28 (mean) | 30 | $3 \times 10^5$ | 16-20 | Whole virion ELISA IgG | 0, 7, 14, 28, 56, 84, 180 |
| Agnandji (2017)[3*] | Phase 1 randomised trial | PACTR201411000919191 | Gabon | 27 (mean) | 10 | $3 \times 10^6$ | 36-39 | Whole virion ELISA IgG | 0, 7, 14, 28, 56, 84, 180 |
| Agnandji (2017)[3*] | Phase 1 randomised trial | PACTR201411000919191 | Gabon | 25 (mean) | 6 | $2 \times 10^7$ | 13-16 | Whole virion ELISA IgG | 0, 7, 14, 28, 56, 84, 180 |
| Agnandji (2016)[4†] | Phase 1 RCT | NCT02287480 | Switzerland | 42 (mean) | 40 | $1 \times 10^7$ | 27-28 | Whole virion ELISA IgG | 0, 14, 28, 180 |
| Agnandji (2016)[4†] | Phase 1 RCT | NCT02287480 | Switzerland | 43 (mean) | 31 | $5 \times 10^7$ | 12-14 | Whole virion ELISA IgG | 0, 14, 28, 180 |
| Boum (2020)[5] | Phase 3, non-randomised trial | PACTR201503001057193 | Guinea | 34.5 (mean) | 26.62 | $2 \times 10^7$ | 90-1124 | GP ELISA IgG, Pseudovirus plaque reduction neutralisation, Whole virion ELISA IgG, Whole virion neutralisation | 0, 14, 28, 180 |
| Davis (2020)[6‡] | Observational study | N/A | UK | 40 (median) | Not reported | $2 \times 10^7$ | 12-17 | GP ELISA IgG, Whole virion neutralisation | 14, 28, 90, 180, 270, 365 |
| ElSherif (2017)[7] | Phase 1 RCT | NCT02374385 | Canada | 34.5 (mean) | 70 | $1 \times 10^5$ | 9-10 | GP ELISA IgG | 0, 7, 14, 28, 56, 84, 180 |
| ElSherif (2017)[7] | Phase 1 RCT | NCT02374385 | Canada | 35.8 (mean) | 50 | $5 \times 10^5$ | 10 | GP ELISA IgG | 0, 7, 14, 28, 56, 84, 180 |
| ElSherif (2017)[7] | Phase 1 RCT | NCT02374385 | Canada | 36.5 (mean) | 60 | $3 \times 10^6$ | 9-10 | GP ELISA IgG | 0, 7, 14, 28, 56, 84, 180 |
| Heppner (2017)[8] | Phase 1b RCT | NCT02314923 | USA | 36.5 (mean) | 50.0 | $3 \times 10^3$ | 51-64 | GP ELISA IgG, Pseudovirus plaque reduction neutralisation | 7, 14, 28, 56, 84, 180, 360 |
| Heppner (2017)[8] | Phase 1b RCT | NCT02314923 | USA | 36.4 (mean) | 51.6 | $3 \times 10^4$ | 52-61 | GP ELISA IgG, Pseudovirus plaque reduction neutralisation | 7, 14, 28, 56, 84, 180, 360 |
| Heppner (2017)[8] | Phase 1b RCT | NCT02314923 | USA | 37.3 (mean) | 48.4 | $3 \times 10^5$ | 56-64 | GP ELISA IgG, Pseudovirus plaque reduction neutralisation | 7, 14, 28, 56, 84, 180, 360 |

| | | | | | | | | | |
|---|---|---|---|---|---|---|---|---|---|
| Heppner (2017)[8] | Phase 1b RCT | NCT02314923 | USA | 38.0 (mean) | 48.8 | 3 x $10^{6}$ | 61-83 | GP ELISA IgG, Pseudovirus plaque reduction neutralisation | 7, 14, 28, 56, 84, 180, 360 |
| Heppner (2017)[8] | Phase 1b RCT | NCT02314923 | USA | 37.7 (mean) | 51.1 | 9 x $10^{6}$ | 38-45 | GP ELISA IgG, Pseudovirus plaque reduction neutralisation | 7, 14, 28, 56, 180, 360 |
| Heppner (2017)[8] | Phase 1b RCT | NCT02314923 | USA | 38 (mean) | 46.8 | 2 x $10^{7}$ | 39-46 | GP ELISA IgG, Pseudovirus plaque reduction neutralisation | 7, 14, 28, 56, 180, 360 |
| Heppner (2017)[8] | Phase 1b RCT | NCT02314923 | USA | 35.9 (mean) | 31.3 | 1 x $10^{8}$ | 41-45 | GP ELISA IgG, Pseudovirus plaque reduction neutralisation | 7, 14, 28, 56, 180, 360 |
| Huttner (2015)[9] | Phase 1/2 RCT | NCT02287480 | Switzerland | 40 (mean) | 53 | 3 x $10^{5}$ | 51 | GP ELISA IgG, Whole virion ELISA IgG | 0, 14, 28 |
| Huttner (2015)[9] | Phase 1/2 RCT | NCT02287480 | Switzerland | 42 (mean) | 40 | 1 x $10^{7}$ | 34-35 | GP ELISA IgG | 0, 14, 28 |
| Huttner (2015)[9] | Phase 1/2 RCT | NCT02287480 | Switzerland | 43 (mean) | 31 | 5 x $10^{7}$ | 13-16 | GP ELISA IgG | 0, 14, 28 |
| Kennedy (2017)[10] | Phase 2 RCT | NCT02344407 | Liberia | 29 (median) | 37.4 | 2 x $10^{7}$ | 485-495 | GP ELISA IgG | 7, 30, 180, 365 |
| Kieh (2022)[11] | Phase 2 RCT | NCT02876328 | Guinea, Liberia, Mali, Sierra Leone | 27 (median) | 46 | 9.4 x $10^{7}$ | 361-388 | GP ELISA IgG | 0, 14, 28, 56, 63, 90, 180, 365 |
| Kieh (2022)[11]¶ | Phase 2 RCT | NCT02876328 | Guinea, Liberia, Mali, Sierra Leone | 26 (median) | 44 | 9.4 x $10^{7}$ | 182-197 | GP ELISA IgG | 0, 14, 28, 56, 63, 90, 180, 365 |
| Kieh (2022)[11] | Phase 2 RCT | NCT02876328 | Guinea, Liberia, Mali, Sierra Leone | 9 (median) | 45 | 9.4 x $10^{7}$ | 391-401 | GP ELISA IgG | 0, 14, 28, 56, 63, 90, 180, 365 |
| Kieh (2022)[11]¶ | Phase 2 RCT | NCT02876328 | Guinea, Liberia, Mali, Sierra Leone | 8 (median) | 42 | 9.4 x $10^{7}$ | 188-200 | GP ELISA IgG | 0, 14, 28, 56, 63, 90, 180, 365 |
| Regules (2017)[12]Δ | Phase 1 trials | NCT02269423/ NCT02280408 | USA | 36.9 (mean) | 35 | 3 x $10^{6}$ | 9-20 | GP ELISA IgG, Pseudovirus neutralisation | 0, 7, 14, 28, 56, 84,180 |
| Regules (2017)[12]Δ | Phase 1 trials | NCT02269423/ NCT02280408 | USA | 34.6 (mean) | 20 | 2 x $10^{7}$ | 9-20 | GP ELISA IgG, Pseudovirus neutralisation | 0, 7, 14, 28, 56, 84,180 |
| Regules (2017)[12]Δ | Phase 1 trials | NCT02269423/ NCT02280408 | USA | 38.8 (mean) | 25 | 1 x $10^{8}$ | 10-20 | GP ELISA IgG, Pseudovirus neutralisation | 0, 7, 14, 28, 56, 84,180 |

* Three cohorts in Agnandji (2017)[3] were excluded as they did not report antibody levels prior to day 28 (adults who received 3 x $10^{5}$ PFU, children and adolescents).

† Only results from Geneva cohort in Agnandji (2016)[4] were included. All other cohorts were excluded as they did not report antibody data before day 28.

‡ Davis (2020)[6] also reported results of a GP ELISA IgM assay for the same cohort. This was excluded as the antibody levels were reported as optical densities.

¶ These cohorts were included in the analysis of fold-change to day 28 but were excluded from the analysis of antibody levels over time as these people received a second dose on day 56.

∆ Timepoints up to day 28 for these cohorts contained pooled results including people who received a second dose on day 28. All timepoints after day 28 that were included in the analysis only include data from people who received a single dose.

**Table S2. Studies identified in the systematic review but not included in the analysis**

| Author (year) | Study design | Trial number | Reason for exclusion |
|---|---|---|---|
| Atre (2019)[13] | Phase 1 RCT | NCT02269423 | Duplicate data reported in other studies |
| Chandrasekaran (2024)[14] | Phase 2 RCT | NCT02344407 | Duplicate data reported in other studies |
| Dahlke (2017)[15] | Phase 1 trial | NCT02283099 | Outcome measurement incompatible with analysis: results reported exclusively as optical density (OD) |
| Grais (2021)[16] | Phase 2/3 trials | PACTR201503001057193/ NCT02344407/ NCT02378753 | Duplicate data reported in other studies |
| Beavogui (2026)[17] | Phase 2 RCT | NCT02876328 | Duplicate data reported in other studies |
| Vianello (2023)[18] | Phase 1b RCT | NCT02314923 | Duplicate data reported in other studies |
| Wiedemann (2024)[19] | Phase 2 RCT | NCT02876328 | Duplicate data reported in other studies |

**Table S3. Fold-change estimates between day 7 or 14 and day 28 using all data compared with restricting to include data on the standard doses only**

| | All data | Standard dose* |
|---|---|---|
| **Day 7 - 28** | **Fold-change (95% CI)** | **Fold-change (95% CI)** |
| Binding | 17.5 (5.3 - 57.4) | 19.8 (4.7 - 83.6) |
| Neutralising | 11.0 (3.0 - 40.9) | 13.4 (2.7 - 67.6) |
| **Random Effects** | | |
| Level 2 $I^2$ | 5.7% | 10.93% |
| Level 3 $I^2$ | 91.6% | 86.66% |
| **Day 14 - 28** | | |
| Binding | 2.6 (1.6 - 4.1) | 2.4 (1.5 - 3.8) |
| Neutralising | 2.1 (1.2 - 3.9) | 2.0 (1.1 - 3.4) |
| **Random Effects** | | |
| Level 2 $I^2$ | 36.83% | 39.67% |
| Level 3 $I^2$ | 59.68% | 57.29% |

* Standard dose is defined as Ervebo doses between $2 \times 10^7$ and $1 \times 10^8$ pfu
